# Natural disease progression vs. DBS-related worsening in essential tremor

**DOI:** 10.64898/2025.12.17.25342504

**Authors:** Nisrine Bakri, Adolfo Ramirez-Zamora, Michael S Okun, Kelly Foote, Evangelos Christou, Karim Oweiss

## Abstract

Essential tremor is a disabling and highly prevalent movement disorder. Deep brain stimulation of the ventral intermediate nucleus provides substantial symptom relief; however, many patients experience diminishing benefit over time. The underlying causes range from natural disease progression and worsening ataxia to, less commonly, habituation or tolerance.

We performed a retrospective longitudinal analysis of 86 individuals with essential tremor who underwent unilateral ventral intermediate nucleus deep brain stimulation and had three or more follow-up visits (mean follow-up duration 5.2 years; maximum 15.4 years; >470 total visits). This approach provided a unique opportunity to assess contralateral limbs in addition to long term effects of stimulation. Tremor severity was quantified using the Fahn–Tolosa–Marin Tremor Rating Scale total score and subscores, with particular focus on contralateral and ipsilateral upper-extremity tremor. Longitudinal trajectories were assessed across limbs and stimulation states at both the population and individual levels. Comparison of tremor between the non-stimulated side and the stimulated side were conducted in the stimulation off condition and served as an intrinsic control to disentangle natural disease progression from tolerance to deep brain stimulation.

Contralateral and ipsilateral upper-extremity tremor assessed with stimulation off showed slow, bilateral progression over time. Tremor in the ipsilateral limb followed the same trajectory as the unstimulated contralateral limb, consistent with natural disease progression. Stimulation exerted no measurable cross-hemispheric influence, as ipsilateral tremor progressed similarly with stimulation on and off. During chronic stimulation, tremor progression in the stimulated limb closely tracked that of the ipsilateral limb, with most patients showing no meaningful difference between sides. Progression in the stimulated limb was also similar during stimulation on and off conditions, suggesting that habituation, stimulation-induced maladaptation, or deep brain stimulation-accelerated decline was not observed in this cohort.

Long-term symptoms of worsening tremor in essential tremor patients with unilateral ventral intermediate nucleus deep brain stimulation were primarily driven by natural disease progression, not by deep brain stimulation-related causes. While tolerance may rarely emerge, the dominant effect of worsening is disease progression. In this cohort, the stimulation programming parameters were individually optimised over time, likely minimizing the influence of suboptimal programming or stimulation-related decline. These findings underscore the importance of extended comprehensive outcome assessment in essential tremor, incorporating objective measures to distinguish disease progression from treatment-related effects. Follow-up studies are needed to better characterise the relative contributions of tremor and ataxia to long-term disability.

## Introduction

Essential tremor (ET) is the most common movement disorder and affects an estimated 25 million people worldwide.^1,2^ ET primarily manifests as a tremor in the upper extremities during goal-directed behaviour, with possible later manifestations in the head, voice, or lower limbs, and without other neurological signs such as ataxia, dystonia, or parkinsonism.^3,4^ ET is however also considered an ataxia syndrome as other symptoms including gait and balance dysfunction emerge later in the disease.^5,6^ ET is usually defined clinically rather than by a single cause, and its underlying pathophysiology, particularly the involvement of the cerebellar-thalamic-cortical network.^3^ Although often characterised by slow progression, accumulating evidence suggests that ET can exhibit more rapid progression over time. Many patients experience worsening tremor amplitude, spread to additional body regions, or increasing functional impairment, underscoring that ET is not uniformly static but can evolve in severity and impact.^7–9^ ET has also been shown to be associated with substantial fall-related morbidity. ^10–13^ The natural trajectory of ET introduces challenges in long–term management especially in distinguishing the underlying course of the disease from treatment-related changes.

Medications including propranolol and primidone and antiepileptics continue to serve as the primary therapeutic approach, yet many patients experience limited symptomatic relief either alone or in combination with considerable side effects.^14^ For medication-refractory cases, procedures including deep brain stimulation (DBS) and MRI-guided focused ultrasound have been shown to improve outcomes.^15–17^ Yet even with effective therapy, concerns persist regarding stimulation-induced side effects, habituation, and the long-term durability of tremor control.^18^ A considerable proportion of patients continue to experience disabling tremor or ataxia that remain insufficiently controlled by DBS alone or worsens over follow-up.

DBS of the ventral intermediate nucleus (VIM), and increasingly the posterior subthalamic area (PSA), has been shown to be among the most effective treatment options, providing substantial tremor suppression.^19–23^ While many patients experience robust and durable tremor suppression, a substantial subset exhibit a gradual reduction in benefit over months to years.^24,25^ The literature has defined these cases by employing heterogeneous terminology (“habituation,” “tolerance,” “therapy escape,” “rebound”) to describe DBS-related effects, reflecting the uncertainty about whether declining efficacy reflects biological adaptation to chronic stimulation, network maladaptation, suboptimal lead placement, or possibly progressive cerebellar or neural network pathology.^26–32^ Emerging evidence points to cerebellar involvement as a consistent contributor: stimulation can modulate parallel cerebellar output pathways including the dentato-rubro-thalamic pathway suppressing tremor at therapeutic intensities, but provoking gait disturbance, dysarthria, or limb ataxia especially when off-target fibers are recruited^33^.

This duality creates a fundamental challenge for the field. The worsening of tremor years after DBS lead implantation raises a critical question: is this decline driven by intrinsic disease progression, stimulation-induced cerebellar dysfunction, habituation or tolerance to chronic stimulation, gradual loss of effective current delivery, or a combination of these mechanisms? Disentangling these mechanisms as well as understanding why some patients maintain long-term benefit while others worsen despite aggressive reprogramming, represents a major unmet need. Addressing this uncertainty will be crucial, not only for prognosis and patient counseling, but also for optimising programming strategies, designing adaptive stimulation paradigms, and for clarifying the mechanistic basis for long-term ET DBS failure.^34–37^

In the present experiment, we aimed to disentangle two key contributors to long-term tremor worsening in essential tremor (ET): intrinsic disease progression, reflecting bilateral, time-dependent changes within the cerebello-thalamo-cortical network, and mechanisms potentially driven by chronic deep brain stimulation itself. Unlike prior studies that have relied on comparisons between separate treated and untreated cohorts, which potentially mix these mechanisms, our approach leveraged within-subject control (stable internal reference) through capitalizing on the lateralized nature of unilateral VIM DBS. Patients implanted unilaterally provided a unique opportunity to use one hemisphere as treated, while using the contralateral brain or non-stimulated hemisphere as an internal control. This design enabled us to examine whether tremor severity on the untreated side progressed independently, whether cross-hemispheric or network-level effects of stimulation influenced the non-stimulated side, and whether the rate of decline differed between sides. We aimed to characterise lateralization and to quantify potential crossover effects. We analysed two complementary control conditions: (i) the non-stimulated side as a naturalistic index of disease progression, and (ii) the stimulated side during DBS-OFF states as a therapy-free representation of the same neural circuit. By tracking longitudinal changes across these conditions, we assessed the trajectory and rate of symptom worsening and elucidated a mechanistically informed framework to separate true disease progression from stimulation-related phenomena.

## Methods

### Patient cohort

We conducted a retrospective cohort study, approved by the University of Florida Institutional Review Board (IRB 201901807) drawing on longitudinal clinical data from the UF INFORM database, a prospectively maintained repository of demographic, clinical, and surgical information for patients evaluated and treated in the Movement Disorders and Neurorestoration Program. All individuals diagnosed with ET who underwent deep brain stimulation (DBS) surgery at the University of Florida between January 2003 and September 2020 were screened, yielding an initial cohort of 137 patients.

Patients were eligible for inclusion if they carried a diagnosis of ET established by a fellowship-trained movement disorders neurologist, had undergone unilateral implantation of a VIM-DBS system at UF, and demonstrated a stable clinical response for at least three months following surgery. Baseline Fahn-Tolosa-Marin Tremor Rating Scale (FTM-TRS) assessments had to be available prior to implantation to permit baseline and longitudinal comparison. Individuals who underwent staged bilateral DBS on the contralateral side or who had fewer than three postoperative follow-up visits were excluded to ensure adequate characterisation of long-term outcomes. A total of 86 patients met the inclusion criteria, contributing more than 470 visits, which comprised the final analysis cohort.

### Assessment of clinical outcomes

The primary outcome of this study was to quantify the longitudinal effectiveness of unilateral VIM DBS by comparing baseline (pre-DBS) total score and subscores of FTM TRS with follow-up scores obtained during stimulation (FTM TRS-ON). As part of routine clinical care, patients undergo standardized follow-up visits at approximately 6 and 12 months after implantation and annually thereafter. At each visit, the FTM TRS are recorded in both stimulation-ON and stimulation-OFF conditions, with a consistent 30-minute washout period preceding the OFF assessment to minimize residual carryover effects.

The FTM TRS is a validated instrument with strong interrater reliability after appropriate training. It consists of three components that capture distinct aspects of tremor severity and functional impact. Part A (items 1-9) and Part B (items 10-14) are fully examiner-rated and evaluate tremor amplitude, laterality, and motor performance across tasks such as handwriting, spiral drawing, and pouring. Part C (items 15-21) comprises patient-reported (subjective) assessments of functional disability in daily activities. To characterise lateralized tremor severity, we also derived right- and left-sided subscores using established item groupings (right: items 5, 8, 11R-14R; left: items 6, 9, 11L-14L)^38,39^.

Secondary analyses focused on separating disease progression from stimulation-related effects by examining longitudinal trajectories of tremor severity across limbs and stimulation states. We first compared contralateral (CUE) and ipsilateral (IUE) tremor scores in the DBS-OFF condition to evaluate whether both limbs exhibited similar rates of worsening over time, thereby establishing the untreated limb as a proxy for natural disease progression. We then assessed whether stimulation altered ipsilateral tremor by comparing IUE scores in DBS-ON and DBS-OFF states; similar slopes in these conditions would indicate that unilateral DBS does not influence tremor in the non-stimulated limb. To investigate whether stimulation-related mechanisms contribute to long-term worsening, we compared CUE and IUE progression during chronic DBS-ON. Divergence between CUE and IUE slopes, or faster worsening of CUE despite active stimulation, was interpreted as evidence of stimulation-related processes such as maladaptive cerebellar responses, habituation, or tolerance. Finally, we compared the progression slopes of the stimulated limb (CUE) during DBS-ON and DBS-OFF assessments. This allowed us to determine whether chronic DBS also alters the natural rate of tremor worsening in that limb or whether both states follow the same disease-driven trajectory. These analyses were performed using smoothed population curves, paired per-patient slope comparisons, and cubic mixed-effects modelling to characterise long-term trajectories with appropriate handling of repeated measures.

### Statistical analysis

All analyses were performed in MATLAB using custom code for longitudinal modelling, visualisation, and mixed-effects regression. To characterise long-term changes in tremor severity, we modelled population-level trajectories using linear and cubic mixed-effects models with random intercepts, allowing for unequal numbers of visits per patient and variable follow-up durations. Smoothed mean ± SEM curves were generated to visualize temporal trends and reduce visit-to-visit variability. Individual patient slopes were extracted from linear fits to enable paired comparisons across limbs (CUE vs IUE) and stimulation states (DBS-ON vs DBS-OFF), with slope differences used to determine whether progression rates diverged within subjects. Percent tremor improvement relative to baseline was quantified at each follow-up visit, and population-level changes in improvement were estimated using regression of percent improvement versus years since implantation. Additional analyses evaluated the distribution of clinical responders (baseline vs first postoperative visit; baseline vs last available visit), the evolution of clinician-rated subdomains and patient-reported functional items over time, and the relationship between subjective disability scores and objective motor ratings using cross-sectional regression.

For DBS-ON longitudinal analyses using non-linear mixed-effects models, time since pre-operative baseline was treated as a continuous variable, with baseline (time = 0) as the reference. Population mean trajectories and 95% confidence intervals were derived from the fitted models, and differences from baseline were assessed using model-based contrasts (*P* < 0.05). Time-varying progression was evaluated using the first derivative of the fitted population mean trajectory, classifying intervals as improving, stable, or worsening based on model-derived uncertainty.

## Results

### Longitudinal dynamics of DBS effectiveness over time

To evaluate whether the therapeutic effect of unilateral VIM-DBS diminishes over time, longitudinal analyses were performed using both population-level and individual patient data. Although some patients have follow-up durations of more than 15 years, group-level analyses were limited to the first 10 years due to increasing data sparsity and attrition. As shown in Fig. 2A, patient numbers declined steadily, dropping below 15 by month 120. At that point, the 95% confidence interval for the mixed-effects model exceeded 14.2 percentage points, over 1.5× the minimum width deemed acceptable for reliable population-level interpretation. Beyond 10 years, analyses were restricted to the individual patient level.

The study cohort included 86 patients (Table 1). The mean age at surgery was 70.3 ± 9.0 years, with a mean follow-up duration of 5.2 ± 3.2 years and a median of 9 visits per patient. Most patients were male (67.4%), with left-sided implantation (83.7%) and right-hand dominance (84.9%). All electrodes targeted the VIM of the thalamus. The mean baseline total FTM TRS score was 50.9 ± 14.1, including a clinician-rated objective score of 35.7 ± 10.7. Contralateral (CUE) and ipsilateral (IUE) upper extremity subscores were 15.7 ± 4.6 and 13.2 ± 5.0, respectively, with a difference between subscores (*P*< 0.001), consistent with greater tremor severity on the planned stimulation side. Baseline global function and handwriting scores were 15.4 ± 4.6 and 2.4 ± 1.1, respectively. A primary diagnosis of ET was present in 89.5% of patients; the remainder had additional etiologies.

**Table 1.** Comprehensive summary of patient demographics and baseline total and subscale assessments.

|  | **Variable** | **Value** |
| --- | --- | --- |
| **Cohort** | N (patients) | 86 |
| **Cohort** | Age at surgery (years) | 70.3 ± 9.0 |
| **Cohort** | Follow-up (years) | 5.2 ± 3.2 |
| **Cohort** | Visits per patient (median [IQR]) | 9 [6–14] |
| **Cohort** | Sex (n, %) | M: 58 (67.4%), F: 28 (32.6%) |
| **Cohort** | Implanted side (n, %) | Left: 72 (83.7%), Right: 14 (16.3%) |
| **Cohort** | Handedness (n, %) | R: 73 (84.9%), L: 11 (12.8%), NULL: 2 (2.3%) |
| **Cohort** | Target (n, %) | Vim: 86 (100.0%) |
| **Cohort** | Diagnosis category (n, %) | ET only: 77 (89.5%), ET + other diagnosis: 9 (10.5%) |
| **Baseline** | Baseline TRS Total (clinician only) | 35.7 ± 10.7 |
| **Baseline** | Baseline TRS Total | 50.9 ± 14.1 |
| **Baseline** | Baseline CUE TRS | 15.7 ± 4.6 |
| **Baseline** | Baseline IUE TRS | 13.2 ± 5.0 |
| **Baseline** | Baseline Global Function | 15.4 ± 4.6 |
| **Baseline** | Baseline Handwriting | 2.4 ± 1.1 |
| **Baseline** | Baseline CUE vs IUE (paired t-test, p) | p = 0.000 |
| **DBS State** | Baseline rows identified (n) | 86 |

Year-by-year percent improvement in total tremor score (DBS-ON vs. baseline) was assessed. We observed a robust initial response during the first year (median ∼70%), with a progressive decline over time (Fig. 2B). Median improvement remained above 50% through year 6, followed by a steeper decline in later years. By years 9–10, the median improvement had fallen below 15%, with improvement remaining different from baseline (*P*< 0.05). Despite this trend, clinically meaningful benefit was retained in a substantial proportion of patients for several years post-implantation.

To quantify the overall decline, a linear mixed-effects model was applied to 470 visits across the full cohort (Fig. 2C). The analysis demonstrated a negative slope, corresponding to an average annual decline in improvement of 3.57% per year (*P* < 0.001). While individual variability was observed, the modelled trend indicates a gradual reduction in DBS effectiveness over time.

To capture non-linear, time-dependent changes in DBS-ON tremor severity, we next applied a longitudinal mixed-effects model with non-linear time terms to total FTM TRS scores. This analysis revealed a time-dependent pattern of initial improvement followed by gradual worsening (Supplementary Fig. 2). Individual patient trajectories demonstrated substantial inter-subject variability, yet the population-average trajectory showed a pronounced reduction in tremor severity after DBS implantation, with sustained benefit during early follow-up (Supplementary Fig. 2A). Population mean estimates remained significantly improved relative to baseline for an extended period, before progressively converging toward pre-operative tremor severity at later time points (Supplementary Fig. 2B). Analysis of the time-varying slope of the population mean trajectory demonstrated a prolonged interval of significantly negative slopes, consistent with continued improvement, followed by a transition to non-significant slopes (indicating a return toward pre-operative tremor severity) and, at later times, significantly positive slopes indicating worsening (Supplementary Fig. 2C).

At the first visit, 86% of patients showed clear improvement in clinician-rated FTM TRS scores, with only 14% classified as non-improved (Fig. 2D). By the last visit (visit ≥6), the proportion of non-improved patients increased to 45.2%, while only 54.8% maintained a net benefit compared to baseline. This shift highlights a progressive decline in individual-level improvement over time, despite initial success early after surgery.

We then analysed FTM TRS subscores across follow-up visits numbers (Fig. 2E). These included clinician-rated scores, contralateral (CUE) and ipsilateral (IUE) tremor scores, global function, and handwriting. Most subscores improved early after surgery but gradually worsened in later visits. By the final follow-ups, several scores had returned close to baseline, creating the impression that the DBS effect had worn off over time, around visit 15 (on average, 85 months post-surgery). However, the IUE score showed a steady increase across visits, consistent with ongoing disease progression.

We then analysed FTM TRS subscores across time. The clinician-rated FTM TRS domains, including axial symptoms, handwriting, and upper/lower extremities on both sides, were analysed through a radar plot (Fig. 2F). Domains typically influenced by stimulation showed marked improvement in the early years. These effects gradually diminished over time, with many domains returning toward baseline by years 7 to 8.

As expected, the IUE showed little initial improvement, since it was not directly targeted by stimulation. However, it worsened steadily over time, consistent with disease progression. The radar plot becomes more rounded in later years, reflecting a general convergence of all domains back toward baseline levels.

Early improvement was also reported across patient-reported activities in speaking, feeding (solids), bringing food to mouth, hygiene, dressing, writing, and working (Fig. 2G). By years 7-10, scores worsened again in nearly all domains, suggesting a decline in benefit over time and consistent with the pattern seen in objective ratings. To quantify this alignment, subjective and objective scores were directly compared. At the population level, subjective scores were positively correlated with clinician-rated FTM TRS scores (Fig. 2H), but the slope was modest (0.38), indicating that patients rated their disability lower than clinicians did, on average. This relationship was further assessed using a Bland-Altman analysis (Supplementary Fig. 1C), which showed that subjective scores were often lower than objective scores, with widespread differences^40,41^. Most differences clustered around a negative mean bias, again reflecting a general underestimation of tremor severity by patients.

To test for outcome differences across clinical subgroups, analyses were conducted based on sex (Fig. 2I), age (Fig. 2J), diagnosis category (essential tremor only vs essential tremor with additional diagnosis) and whether the dominant side was stimulated (treated) or not (Supplementary Fig.1 A and Fig.1 B). No differences in average DBS benefit were observed between males and females (*P* = 0.7273), or across age (slope = 0.35, *P* = 0.340). Patients with ET only showed limited evidence for greater improvement compared to those with an additional diagnosis (*P* = 0.0953). Similarly, stimulation of the dominant side did not influence outcomes (*P* = 0.2928). These results suggest that baseline demographic and clinical variables had limited impact on long-term DBS efficacy in this cohort. Finally, we examined whether longer postoperative follow-up influenced the estimated patient-level slopes in the DBS-ON condition (Fig. 2K). Follow-up duration was positively correlated with individual DBS-ON slopes (*r* = 0.54, *P* < 0.001), indicating that patients with shorter follow-up tended to show more negative slopes, reflecting stronger early benefit, whereas those followed for longer periods exhibited flatter slopes approaching zero.

**Figure 1.**
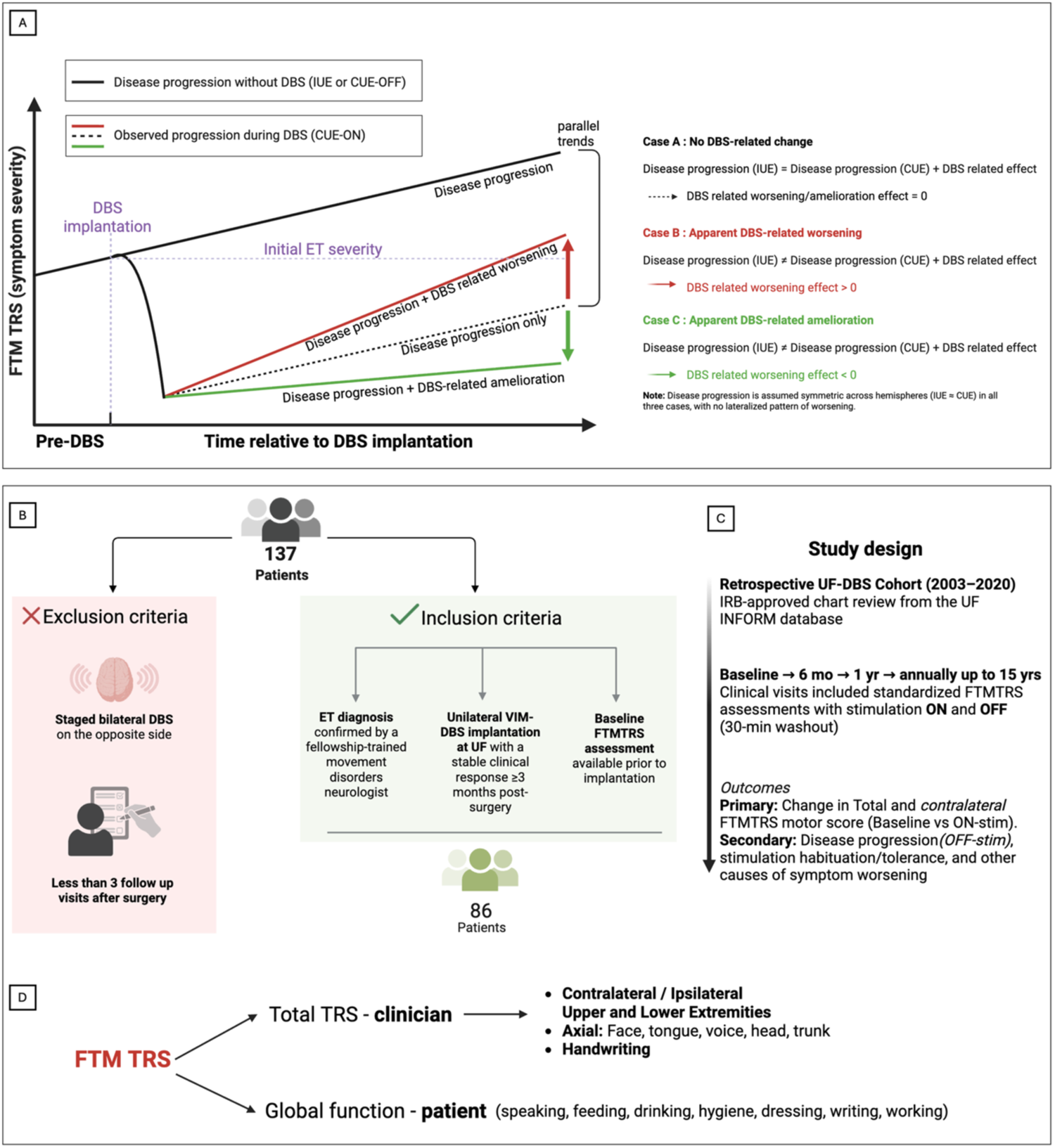
Conceptual framework, cohort selection, and outcome measures for long-term DBS analysis. **(A)** Conceptual schematic illustrating three possible longitudinal relationships between intrinsic disease progression and observed tremor severity during chronic deep brain stimulation (DBS). The black line represents disease progression in the absence of stimulation, inferred from the ipsilateral upper extremity (IUE) or contralateral upper extremity measured in the DBS-OFF state (CUE-OFF). Colored lines represent observed progression during DBS (CUE-ON). Case A depicts no DBS-related change (parallel trajectories). Case B depicts apparent DBS-related worsening (observed progression exceeds disease-only trajectory). Case C depicts apparent DBS-related amelioration (observed progression is attenuated relative to disease-only trajectory). Disease progression is assumed symmetric across hemispheres in all cases. **(B)** Cohort selection. From 137 ET patients who received DBS, 86 met criteria for unilateral VIM with confirmed ET diagnosis, baseline FTM TRS scores, and ≥3 postoperative follow-ups. Patients with staged bilateral DBS were excluded. **(C)** Study design. Retrospective UF-DBS cohort (2003-2020) with baseline, 6-month, 1-year, and annual visits up to 15 years. Each visit included standardized FTM TRS testing in DBS-ON and DBS-OFF (30-min washout). Primary outcome: change in total and contralateral motor FTM TRS. Secondary outcomes: disease progression (OFF), habituation/tolerance, and other contributors to long-term worsening. **(D)** Clinical measures. The FTM TRS includes examiner-rated motor scores (axial, upper/lower extremities) and patient-reported functional disability across daily tasks. These components were used to track objective tremor severity and subjective impact over time.

**Figure 2.**
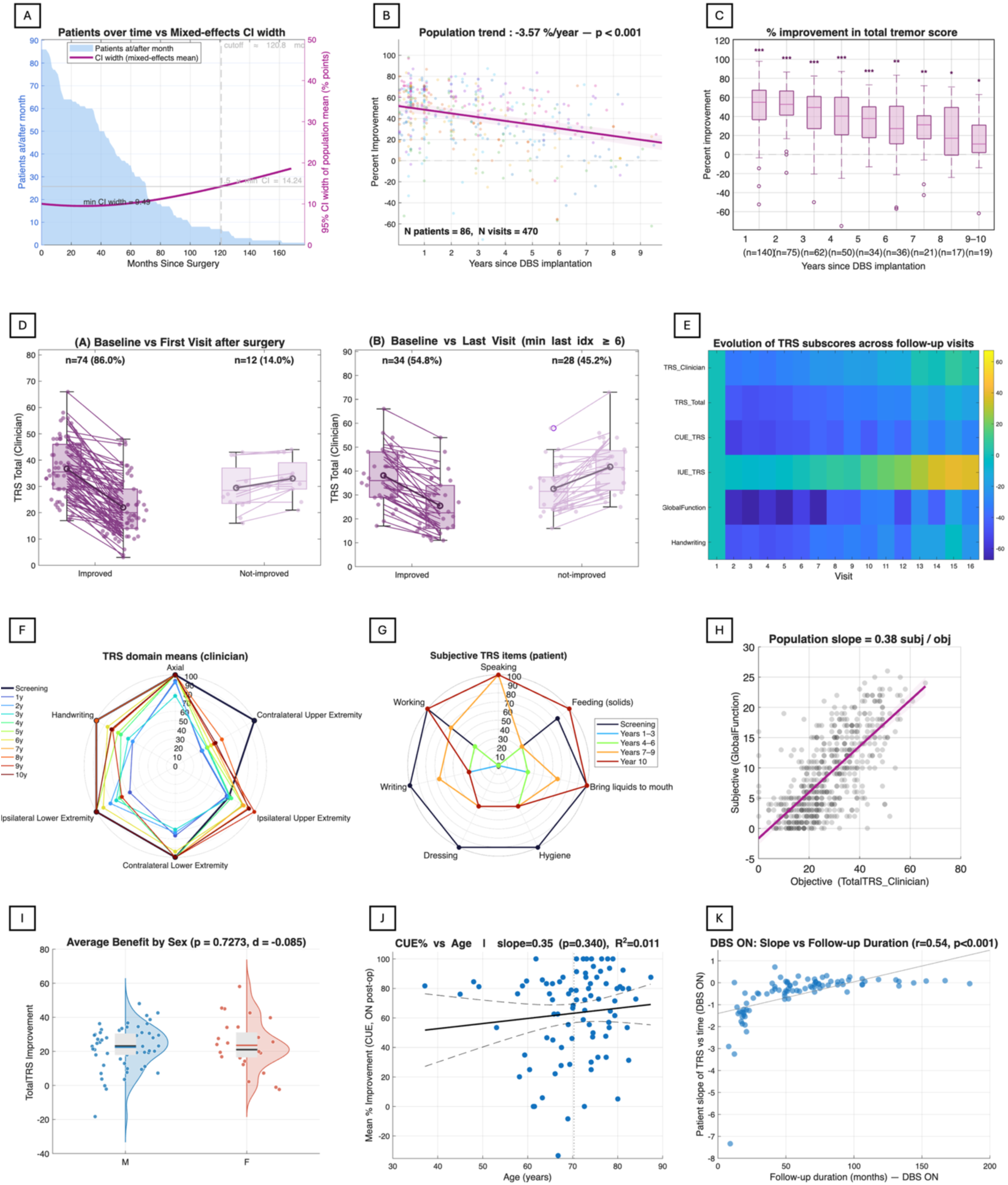
Longitudinal analysis of DBS-treated tremor. **(A)** Number of patients contributing to the data per postoperative month (max 86, tapering to 1 at 160 months) and corresponding mixed-effects 95% confidence interval (CI) width. Precision is highest early after surgery (minimum CI width 0.49 at month 14.2) and widens after patient attrition beyond ∼120 months. **(B)** Percent FTM TRS improvement across 470 visits from 86 patients. Mixed-effects regression shows a decline of −3.57% per year (*P* < 0.001). **(C)** Cross-sectional improvement by postoperative year (*n*=140, 75, 62, 50, 34, 36, 21, 17, 19). Median benefit decreases from ∼58% at year 1 to ∼32% at years 9-10. Significance relative to zero is indicated (*,**,***). **(D)** Heatmap of domain-specific FTM TRS evolution. Contralateral (CUE) tremor shows the greatest and most durable improvement (up to −60%), while ipsilateral (IUE) tremor progressively worsens (+20-60%) over long-term follow-up. **(E)** Paired changes vs baseline. First postoperative visit: 74 improved (86.0%), 12 not improved; median FTM TRS reduction −23, *P* < 0.001. Last visit (index ≥ 6): 34 improved (54.8%), 28 not improved; median −8, *P =* 0.004. **(F-G)** Radar plots of clinician-rated and patient-reported domains show early postoperative benefit with gradual degradation in axial, handwriting, feeding, and dressing tasks beyond years 7-10. **(H)** Strong association between clinician-rated FTM TRS and subjective global function (slope 0.38, *r* ≈ 0.62, *P* < 0.001). **(I)** No sex difference in benefit (M=24.8%, F=22.3%, *P* = 0.7273, d = −0.085). **(J)** No meaningful age effect on CUE improvement (slope 0.35, *P =* 0.340, *R²* = 0.011). **(K)** Greater follow-up duration correlates with more stable patient-specific slopes (*r* = 0.54, *P* < 0.001).

### Validation of the ipsilateral side as a natural disease progression marker

To determine whether unilateral DBS exerts any measurable influence on the non-stimulated IUE, we compared longitudinal tremor trajectories during DBS-ON to DBS-OFF states (Fig. 3A). Population-level modelling showed nearly identical progression rates across conditions: the IUE worsened at 0.077 FTM TRS points per month in the OFF state and 0.075 FTM TRS points per month in the ON state (Fig. 3B). There was no difference between these slopes (*P* = 0.883), indicating that stimulation does not alter long-term tremor progression in the ipsilateral limb. Individual patient slopes were also highly correlated between ON and OFF states (Pearson *r* = 0.739, Spearman *ρ* = 0.654, both *P* < 0.0001), confirming that patients who progressed faster in DBS-OFF also progressed faster in DBS-ON states. Smoothed mean trajectories supported these findings: the IUE-ON and IUE-OFF curves remained virtually superimposed over nearly a decade of follow-up, with mean scores rising from approximately 12-13 points early after surgery to 18-22 points by year 8-9 (Fig. 3C). These results demonstrate that unilateral VIM-DBS has no detectable effect on tremor severity in the ipsilateral upper extremity, reinforcing that the IUE reflects disease progression without detectable modulation by stimulation^42^.

**Figure 3.**
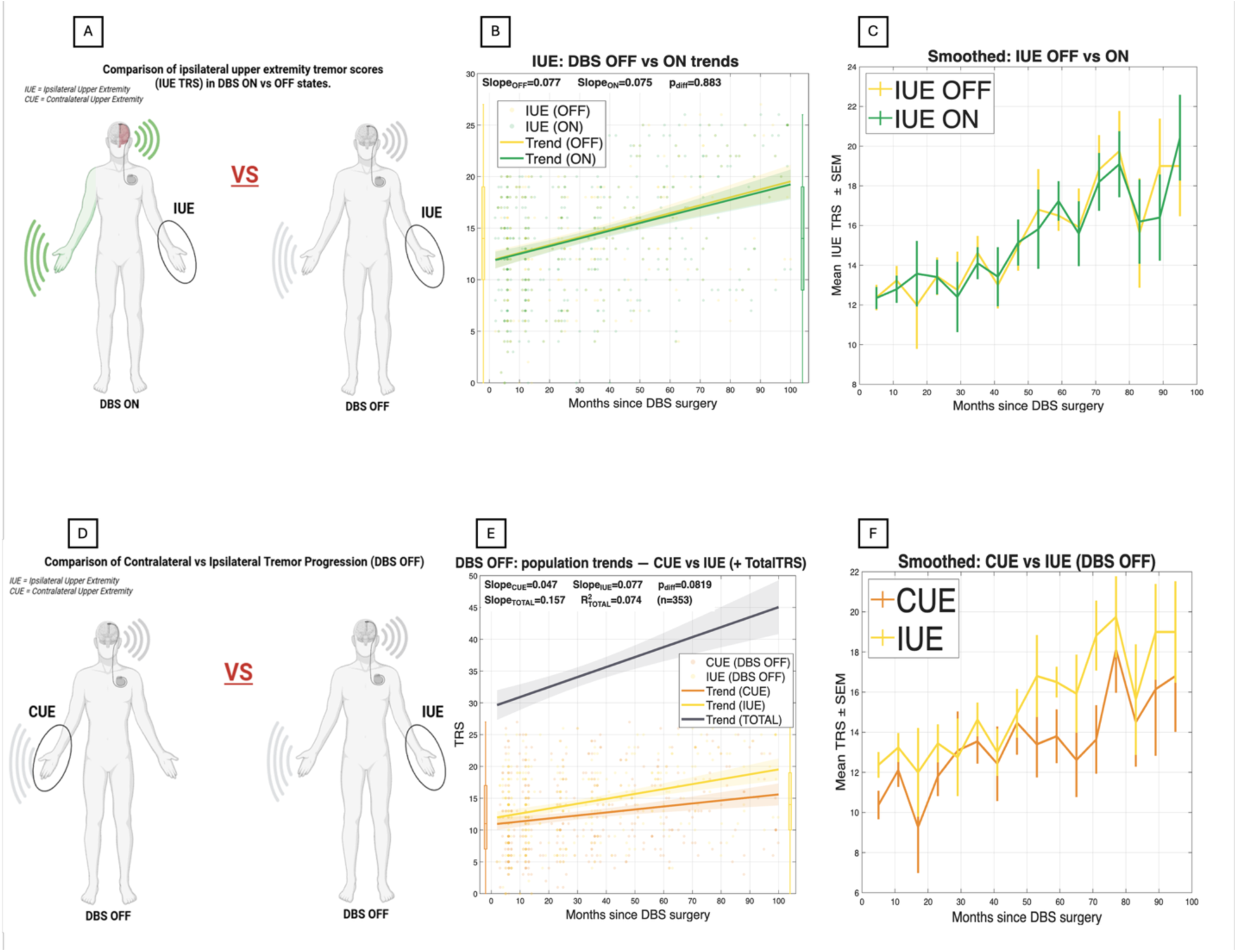
Validation of the ipsilateral side as a natural disease progression marker. **(A)** Schematic comparison of ipsilateral tremor (IUE) during DBS-ON vs OFF conditions, highlighting potential stimulation-related differences. **(B)** Mixed-effects trajectories for IUE in DBS-OFF vs DBS-ON. Slopes were nearly identical (0.077 vs 0.075 FTM TRS/month, *P =* 0.883), indicating that stimulation did not alter long-term ipsilateral tremor progression. Inset shows patient-level slopes (ON vs OFF). **(C)** Smoothed mean IUE trajectories (± SEM) under DBS-OFF and DBS-ON states across postoperative months. Both curves show similar upward trends, consistent with the slope results in B. **(D)** Schematic comparison of contralateral upper extremity (CUE) and ipsilateral upper extremity (IUE) tremor when stimulation is OFF. CUE reflects the stimulated side of the body, whereas IUE represents the non-stimulated limb. **(E)** Mixed-effects population trajectories for CUE and IUE in DBS-OFF. CUE shows a mild progression (slope = +0.07 FTM TRS/month), similar to IUE (slope = +0.07 FTM TRS/month, *P =* 0.8619 for slope difference). Total FTM TRS progression shows a steeper trend (slope = +0.157 FTM TRS/month, *r* = 0.474, *n* = 353). Shaded areas indicate 95% confidence intervals. **(F)** Smoothed mean trajectories.

During DBS-OFF assessments, both the contralateral (CUE) and ipsilateral (IUE) upper extremities showed gradual, measurable worsening in tremor severity over time. Population-level linear modelling demonstrated nearly parallel progression rates between the two limbs, with a CUE slope of 0.047 FTM TRS points per month (≈ 0.56 points/year) and an IUE slope of 0.077 FTM TRS points per month (≈ 0.92 points/year). There was no difference between these slopes (*P* = 0.0819, *n* = 353 DBS-OFF visits), indicating no meaningful divergence in long-term tremor worsening between hemispheres when stimulation is absent. Mixed-effects modelling of total FTM TRS motor scores yielded a comparable overall slope of 0.157 points per month (R² = 0.074), confirming slow but continuous bilateral progression (Fig. 3E).

Smoothed mean trajectories reinforced these findings; although the IUE exhibited slightly higher absolute tremor scores at several time points, both limbs followed overlapping temporal patterns, with mean CUE and IUE scores rising from approximately 11-13 points early after implantation to 15-19 points by 8-9 years post-surgery (Fig. 3F).

Taken together, these findings indicate that there is no lateralised pattern of worsening, and the DBS-OFF stimulated limb (CUE-OFF) can be validly used as an internal control for natural disease progression, equivalent to the ipsilateral limb.

### Disentangling disease progression from stimulation-related effects on contralateral tremor

Having established that the IUE provides a reliable internal marker of natural disease progression, we next examined whether chronic stimulation introduces additional, stimulation-related worsening in the CUE, the limb directly targeted by VIM-DBS. If DBS remains consistently therapeutic over time, CUE progression during stimulation should closely mirror IUE progression. Conversely, any excess CUE worsening would suggest stimulation-related mechanisms such as habituation, maladaptive cerebellar effects, or tolerance (Fig. 1A).

When comparing long-term DBS-ON tremor progression between CUE and IUE, both limbs exhibited slow but steady worsening over years. The CUE progressed at 0.041 FTM TRS points per month, while the IUE progressed at 0.068 FTM TRS points per month. There was no evidence of a difference between these slopes (*P* = 0.0765, *n* = 353 visits), and their 95% confidence intervals showed broad overlap. Thus, at the group level, chronic stimulation did not increase the rate of tremor decline in the stimulated limb relative to the non-stimulated limb (Fig. 4B).

**Figure 4.**
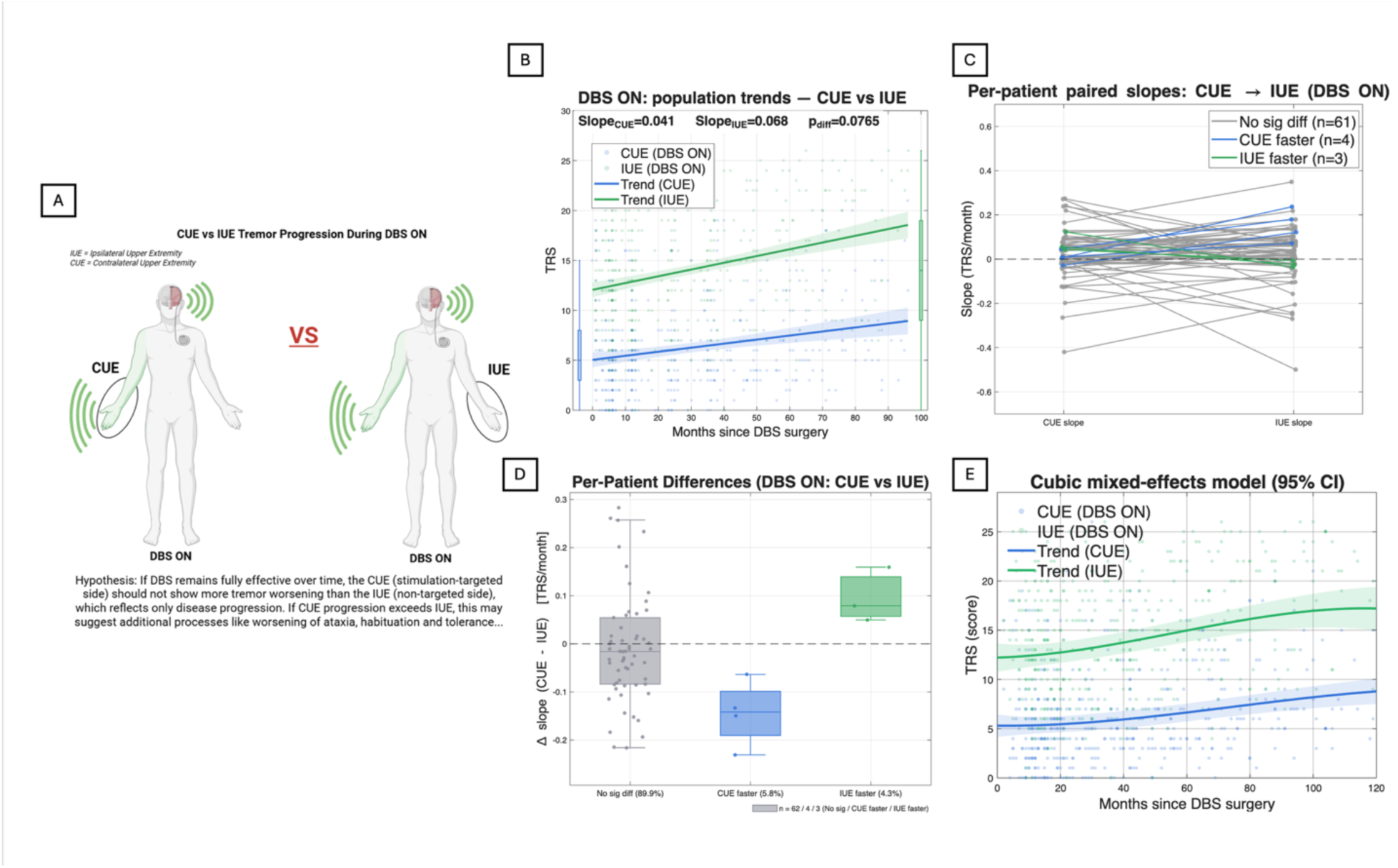
Long-term DBS-ON progression of contralateral (CUE) vs ipsilateral (IUE) tremor. **(A)** Illustration of CUE (stimulated limb) vs IUE (non-stimulated limb) under DBS-ON. If DBS remains effective, CUE should not worsen faster than IUE, which reflects natural disease progression. **(B)** Population mixed-effects trajectories. CUE shows mild progression (0.041 FTM TRS/month) and IUE a slightly faster slope (0.068 FTM TRS/month), with a trend toward difference (*P =* 0.0765). **(C)** Paired within-patient slopes (*n* = 68). Most patients show no difference (61/68, 89.7%); 4 show faster CUE worsening; 3 show faster IUE worsening. **(D)** Distribution of Δ slope (CUE - IUE). Median difference ≈ 0; subgroup proportions match panel C (89.9% no difference; 5.8% CUE faster; 4.3% IUE faster). **(E)** Cubic mixed-effects model (95% CI) showing gradual long-term progression in both limbs, with IUE maintaining a slightly steeper trajectory under DBS-ON.

To avoid masking individual variability, we compared within-subject CUE and IUE slopes for each patient. Strikingly, 61 of 68 patients (89.7%) showed no significant difference between sides. Only 4 patients (5.9%) showed faster CUE worsening, while 3 patients (4.3%) showed faster IUE worsening. This bilateral symmetry reinforces that unilateral DBS does not systematically accelerate or decelerate tremor worsening in the targeted limb (Fig. 4C). The distribution of Δ-slopes (CUE - IUE) centered tightly around zero, with the median approximating 0 FTM TRS points per month, indicating highly symmetric within-brain progression. The few outliers did not cluster in a direction consistent with stimulation-induced deterioration (Fig. 4D).

Finally, cubic mixed-effects curves confirmed that over nearly a decade of follow-up, CUE and IUE trajectories remained parallel, with overlapping 95% confidence bands. Even when allowing for nonlinear changes in slope over time, the stimulated and non-stimulated limbs followed nearly identical long-term paths (Fig. 4E).

### Within-side (CUE) comparison of DBS-ON versus DBS-OFF progression

Building on the preceding analyses showing symmetric bilateral disease progression and minimal stimulation spillover to the ipsilateral limb, we next examined whether chronic DBS alters the long-term trajectory of tremor worsening within the stimulated side itself. In principle, if DBS retains stable long-term efficacy, CUE should worsen at a similar rate in DBS-ON versus DBS-OFF states (Fig. 5A). Conversely, if stimulation-related processes (for example, habituation, stimulation-induced cerebellar dysfunction) accumulate over time, DBS-ON deterioration would exceed the DBS-OFF trajectory.

**Figure 5.**
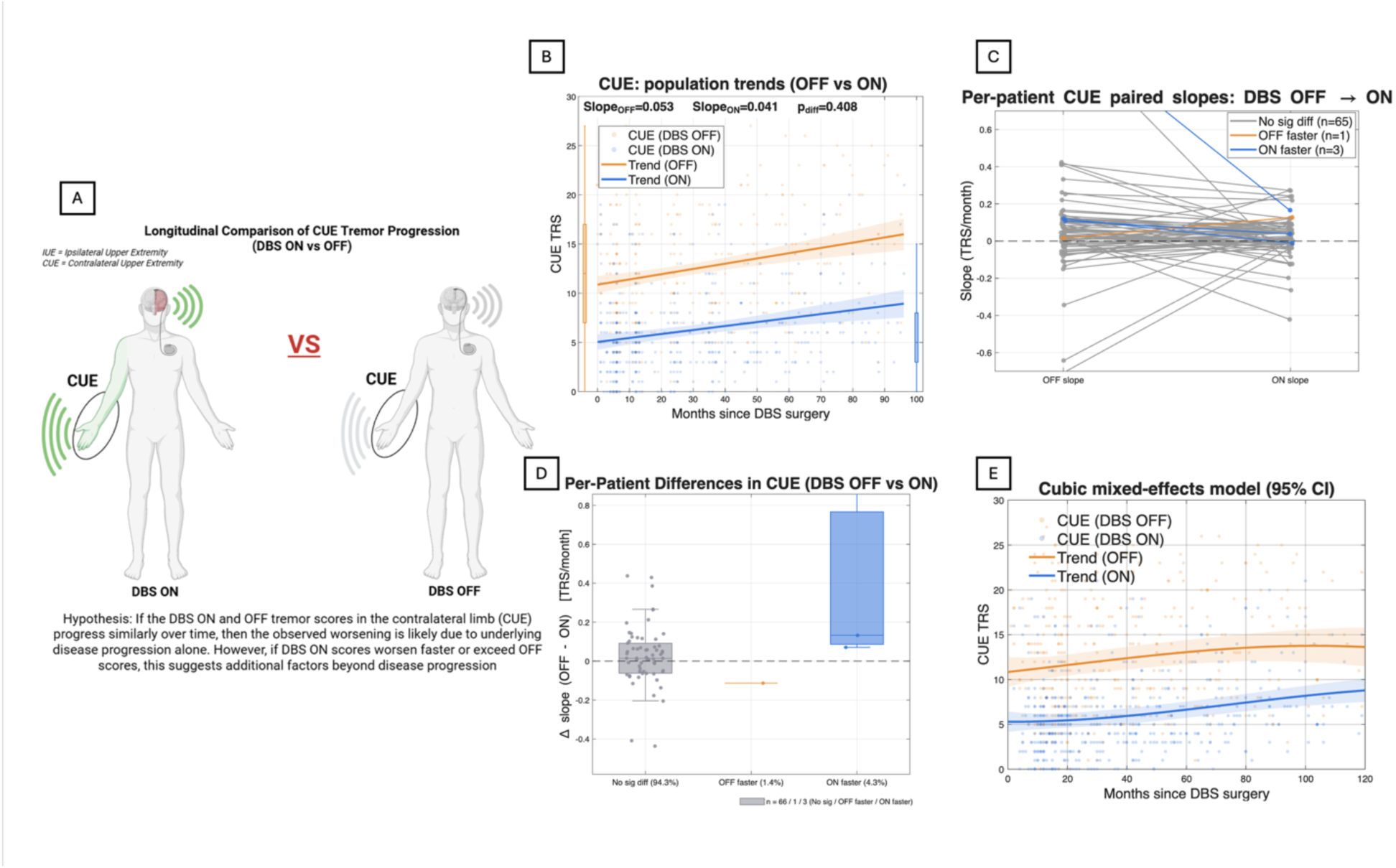
Long-term comparison of contralateral (CUE) tremor progression during DBS-ON vs DBS-OFF. **(A)** Illustration of CUE tremor under DBS-ON vs OFF. If ON and OFF trajectories are similar, worsening likely reflects natural disease progression; greater ON-state worsening may suggest reduced stimulation efficacy or additional mechanisms. **(B)** Population mixed-effects trajectories for CUE in OFF vs ON states. OFF shows a slope of 0.053 FTM TRS/month, ON shows 0.041 FTM TRS/month, with no difference (*P =* 0.408). Shaded areas represent 95% CI. **(C)** Paired within-patient slopes (*n* = 69). Most patients showed no significant OFF→ON slope difference (65/69, 94.2%); 1 had faster OFF worsening; 3 had faster ON worsening. **(D)** Distribution of Δ slope (OFF - ON). Median difference ≈ 0, with subgroup proportions matching panel C (94.3% no difference; 1.4% OFF faster; 4.3% ON faster). **(E)** Cubic mixed-effects model (95% CI) demonstrating similar long-term trajectories in OFF and ON states, with both showing gradual progression consistent with underlying disease.

At the population level (Fig. 5B), the CUE progression slopes were nearly identical across conditions: 0.053 versus 0.041 points per month (*P* = 0.408). Both regression bands rose gradually over time, confirming slow but measurable tremor progression, yet the two curves remained tightly aligned throughout the full follow-up window. Smoothed mean trajectories (Fig. 5C) reinforced this finding: DBS-ON and DBS-OFF curve trajectories were almost parallel, with overlapping SEM envelopes and no divergence at later time points. Per-patient paired-slope comparisons (Fig. 5D) demonstrated the same pattern at the individual level. The vast majority of patients (94.3%) showed no slope difference between DBS-ON and DBS-OFF states. Only 1 patient (1.4%) exhibited faster OFF-state worsening, while 3 patients (4.3%) showed faster ON-state worsening, numbers that are too small to indicate a systematic stimulation-related decline. Together, this strongly argues against large-scale habituation or DBS-induced acceleration of tremor progression in the stimulated limbside. Finally, nonlinear cubic mixed-effects modelling (Fig. 5E) confirmed the robustness of these observations. Both ON and OFF trajectories displayed similar curvature and long-term profiles, with overlapping 95% confidence intervals across nearly the entire follow-up range. No model suggested divergence attributable to chronic stimulation.

## Discussion

### Long-term trajectories of therapeutic benefit and functional decline

The primary goal of this study was to disentangle natural disease progression from stimulation-related causes of long-term tremor worsening in ET patients treated with unilateral VIM-DBS. Using the unstimulated upper extremity and DBS-OFF assessments of the stimulated limb as internal controls, we aimed to determine whether late clinical decline reflected primarily the intrinsic bilateral evolution of the disorder or habituation or alternative stimulation-induced mechanisms.

In this context, the cohort demonstrated a strong initial therapeutic efficacy (tremor suppression) following unilateral VIM-DBS implantation (median ∼70% improvement at year 1) that gradually declined over time, remaining above 50% through mid-term follow-up. By years 9-10, the median improvement had fallen below 15%, while remaining statistically significant. This temporal profile is broadly consistent with prior VIM-DBS series, which typically report 50-80% tremor reduction in the first 1-3 years with gradual loss of efficacy thereafter, but those studies often provide only coarse time points (e.g., “long-term” at 5 or 7 years) and less granular characterisation of the therapeutic trajectory.^43–58^ Except for one study, in which there was year by year follow-up, the sample size was small, starting with a maximum of 14 patients at 2 years and decreasing to 5 patients by year 10 (visits every 3-6 months), likely explaining some lack of statistically significant changes.^59^ In our study, linear mixed effects models were applied across 470 visits, with population level inferences restricted to the first 10 years based on confidence interval width, acknowledging that estimates beyond this window become unreliable because of sample attrition and widening intervals. This data sparsity and model uncertainty contrasts with other long-term ET DBS reports that presented group means at late time points with small sample sizes.^59^ Our approach also provided a precise estimate of the average annual decline in therapeutic efficacy (3.57% per year) and demonstrated that the worsening was slow and progressive, rather than abrupt.

The per-visit analysis in this study reveals the gradual shift in responder distribution and the return of several subscores (e.g., global function, handwriting, contralateral upper extremity) toward baseline by later visits, offering a more nuanced view of how and when functional domains degrade.

Analysis of clinician-rated subscores and patient-reported function shows the expected strong early improvements in stimulation-sensitive domains, followed by gradual convergence of all domains back toward baseline by years 7-8. This “rounding” of the radar plots is conceptually similar to earlier observations that head, voice, and axial tremor may *improve less* and *worsen earlier* than upper limb tremor, however the present work extends that by showing the evolution of handwriting, global function, and other extremity scores over time in the same cohort.^60–62^ The finding that the ipsilateral upper extremity exhibits no initial improvement yet worsens steadily across years is expected, but critical, as it provides an internal comparison and reinforces a progressive, bilateral disease model.

The modest correlation (slope ≈0.38) between subjective disability scores and clinician-rated FTM TRS scores highlights a clinically relevant dissociation. This mirrors findings in Parkinson disease and ET cohorts where patient-reported quality of life and objective motor ratings only partially align, suggesting that mood, adaptation, and expectation substantially modulates perceived benefit.^40,41^ These concerns likely reflect limitations in our current scales and the limited temporal associated with in clinic assessments. In contrast to some earlier ET DBS reports that emphasize concordant improvements in FTM TRS and quality-of-life scales, these data underscore that patients may feel functionally “better” or “worse” than the motor scores suggest, especially in later years when compensatory strategies and acceptance may shift self-assessment. A reasonable criticism is that relying exclusively on the FTM TRS Part C as the subjective measure may miss broader psychosocial dimensions; incorporating disease-specific quality-of-life instruments and mood scales in future work would further clarify how patients internalize long-term DBS benefit.^40,41^

The absence of differences in long-term DBS benefit by sex, age, dominant-side stimulation, or pure ET versus ET-plus other diseases contrasts with earlier reports suggesting that younger age, shorter disease duration, or fewer additional neurologic features might predict better outcomes.^63^ Given the sample size and follow-up distribution, these results imply that such demographic effects are either weaker than assumed or overshadowed by inter-individual variability in disease biology, lead placement, or programming, all of which were not fully modelled here.^64,65^ Thus, the null subgroup findings should not be taken as definitive evidence of no effect but rather highlight that routinely used baseline clinical variables are poor predictors of long-term trajectory and that more specific anatomical and physiological markers (for example, tractography-based targeting) will be likely required for meaningful prognostic stratification. ^64^

### Limited cross-hemispheric influence of unilateral VIM-DBS

Several reports have suggested that unilateral thalamic stimulation may influence tremor bilaterally via network-level or cerebellar mechanisms, supported by imaging and tractography studies showing that both VIM and PSA stimulation engage cerebello-thalamo-cortical pathways and can modulate multiple effectors within this network. Short-term bilateral changes in tremor and physiology have also been described in small series and case reports of unilateral thalamic DBS, pointing to possible cross-hemispheric modulation.^66^

In this cohort, however, the nearly identical IUE-ON and IUE-OFF slopes, together with superimposed smoothed trajectories over almost a decade, indicate that unilateral VIM-DBS does not measurably alter the long-term evolution of tremor in the ipsilateral limb at routine clinical settings. This observation is consistent with larger clinical series comparing unilateral and bilateral VIM/PSA DBS, where robust contralateral tremor control contrasts with limited and often transient effects on the ipsilateral side, ultimately prompting staged implantation when contralateral tremor becomes disabling.^60,67^ Taken together, these data suggest that while VIM-DBS modulates a cerebello-thalamo-cortical network, its durable therapeutic action remains largely confined to the stimulated projection and does not reshape the natural history of the opposite hemisphere, providing a mechanistic rationale for the frequent need for bilateral or staged procedures in ET.^60^

### Parallel Disease Progression in Treated and Untreated Hemispheres

A central question in the long-term evaluation of thalamic DBS for ET is whether the apparent decline in benefit reflects stimulation-related failure or simply the natural evolution of a bilateral disorder. Across decades of published experience, most longitudinal DBS series describe a slow, symmetric increase in tremor severity on both sides of the body, even when only one hemisphere is implanted.^19,48^ Thus, many groups have proposed that ET behaves as a fundamentally bilateral degenerative process, in which untreated regions worsen at a pace that mirrors the stimulated side.^18^ Our data provides strong support for this model. When stimulation was withdrawn, the contralateral and ipsilateral extremities showed parallel, nearly overlapping trajectories of tremor worsening. The progression slopes were statistically indistinguishable, and at no point did the stimulated hemisphere exhibit a unique pattern of decline suggestive of DBS-related harm. This parallelism persisted across nearly a decade of follow-up, despite inter-individual variability in baseline severity and visit frequency. This symmetry indicates that once the acute masking effect of stimulation is removed, both limbs reveal the same underlying disease trajectory. This interpretation reinforces the notion that unilateral DBS neither accelerates nor protects the natural course of the opposite hemisphere.

### Disentangling Disease Progression from Habituation to Stimulation

The distinction between true disease progression and habituation to stimulation, often labeled “tolerance”, remains one of the most contentious issues in the DBS literature. Early work suggested that prolonged, high-intensity stimulation might induce maladaptive plastic changes within tremor circuits, gradually reducing therapeutic benefit. ^68,69^ More recent post-mortem observations, although limited, point toward the possibility of biological adaptation to chronic stimulation.^64,70^ Because both habituation and natural progression can manifest as a decline in DBS efficacy, disentangling the relative contribution of each process is inherently difficult.^70^

Reports in the field differ widely: some groups describing habituation in most patients, up to 73% over roughly 5 years of follow-up, with cases appearing as early as 10 weeks after implantation. ^71^ Conversely, other analyses argue that much of the long-term deterioration seen in ET reflects the underlying course of the disease, rather than stimulation-related tolerance. For example, Favilla et al.^18^ demonstrated comparable tremor worsening in non-DBS ET controls and in patients evaluated with DBS turned both on and off. ^72^ Similarly, a large retrospective cohort followed by Tsuboiet al.^51^ showed that meaningful benefits from VIM DBS can persist for more than a decade in individuals with essential or dystonic tremor. ^73^

This present study provides several lines of convergent evidence that long-term worsening is driven predominantly by disease progression rather than stimulation-related habituation. First, CUE-ON progression is indistinguishable from both IUE-ON and CUE-OFF in most patients, and within-subject slope differences cluster tightly around zero, indicating that the stimulated side does not deteriorate faster than its internal controls. Second, both limbs show slow, bilateral worsening in the OFF state, validating the IUE and CUE-OFF as naturalistic markers of intrinsic progression and demonstrating that the underlying disease advances at a similar rate irrespective of stimulation history. Third, even with follow-up extending beyond a decade and repeated opportunities for maladaptive plasticity to emerge, only a small minority of individuals exhibited asymmetrically greater decline on the stimulated side, and these outliers did not cluster in a manner consistent with a reproducible stimulation-induced phenomenon. The small subset of individuals who deviated from parallel progression patterns may provide insight into how DBS interacts with ongoing disease evolution. In cases where the ipsilateral upper extremity (IUE) appeared to worsen faster than the contralateral, stimulated limb, DBS may be dynamically compensating for newly emerging tremor related to disease progression rather than merely suppressing the original pre-implant symptom burden. In this scenario, stimulation effectively tracks the evolving pathology, maintaining relative symptom control on the treated side while untreated regions reveal the true pace of progression. These individuals represent a clinically informative subgroup that warrants targeted investigation, including detailed review of longitudinal programming history, parameter adjustments, and anatomical factors, to better understand why DBS response diverges from the dominant population pattern.

Taken together, our results indicate that long-term worsening of tremor in ET patients with unilateral VIM-DBS is predominantly explained by the natural progression of the underlying disorder rather than by stimulation-induced habituation. An important aspect of this cohort is that stimulation parameters were systematically reviewed and individually optimised during regular follow-up, with adjustments in amplitude, pulse width, frequency, and active contacts as needed. This procedure reduced the likelihood that the observed decline simply reflected suboptimal programming or a failure to respond to correctable technical issues, and instead supported the interpretation that DBS continues to track a moving target: it effectively suppresses the original, pre-implant tremor burden but cannot fully compensate for additional disease-driven tremor and ataxia that accumulate over time. It should be noted that failure to provide meaningful benefit within the first year is likely related to suboptimal targeting or programming. Additionally, could this visit-by-visit optimization of stimulation parameters be the reason habituation was not clinically apparent in our cohort, thereby allowing DBS to maintain a stable suppression of the original tremor burden by continually updating the stimulation pattern, rather than using a fixed, unchanging program?

In practical terms, these findings suggest that VIM-DBS produces a relatively stable reduction in tremor severity early after implantation, and that later increases in total FTM TRS scores largely reflected new or progressive pathology superimposed on this shifted baseline rather than a true erosion of the initial stimulation effect. This perspective helps reconcile the persistent ON-OFF differences observed years after surgery with the common clinical perception of wearing off, and it underscores the need for long-term outcome measures that extend beyond conventional FTM TRS scoring. Incorporating quantitative kinematic assessments, wearable sensors, brain activity analysis (thalamic LFP) and dedicated ataxia and gait scales will be essential to distinguish progression of tremor from emerging cerebellar dysfunction and to determine whether future strategies, such as adaptive stimulation, smarter contact selection, or targeting of additional nodes within the cerebellar-thalamocortical network, can modify the trajectory of disease-related decline rather than merely masking its earliest manifestations.

To conclude, long-term tremor worsening after unilateral VIM-DBS in ET appears to be driven mainly by the underlying disease course rather than by stimulation-induced habituation or tolerance. Because of clear progressive worsening overtime, clinicians should continually consider the timing for surgery and utilize DBS as soon as functional disability ensures despite adequate medications trials. Unilateral VIM-DBS provides a large early benefit that gradually diminishes as tremor progresses on both the stimulated and non-stimulated sides, with contralateral DBS-ON trajectories closely tracking internal markers of natural progression in almost all patients. These results support using the non-stimulated limb and DBS-OFF assessments as internal controls and suggest that future work should focus on objective tremor and ataxia metrics, circuit-level biomarkers, and advanced stimulation strategies to modify disease-driven decline rather than simply compensating for it.

## Data Availability

The data that support the findings of this study are derived from the University of Florida INFORM database and contain protected health information. Due to ethical restrictions and institutional review board (IRB) regulations, these data are not publicly available. De-identified data supporting the conclusions of this article may be made available from the corresponding author upon reasonable request and subject to institutional approvals and data use agreements.

## Supplementary material

Supplementary material is available at *Brain* online.

## Supplementary material

**Supplementary Figure 1.**
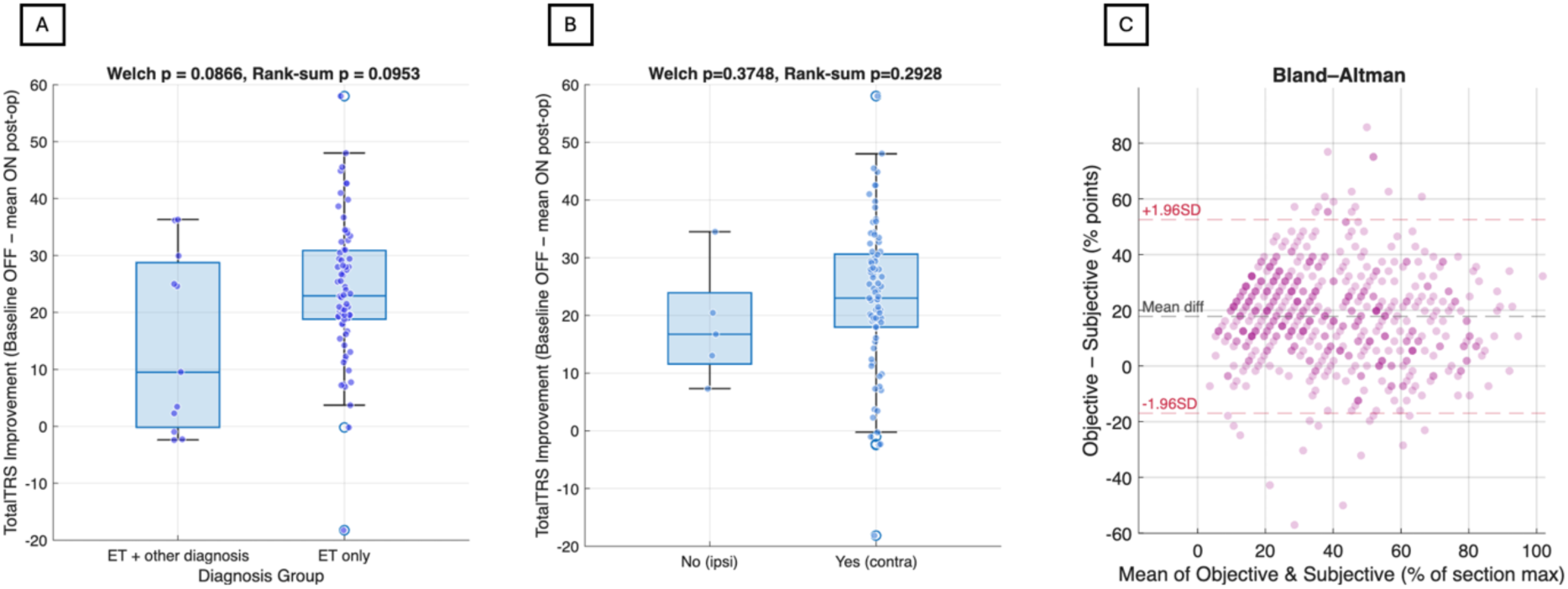
Subgroup analyses of DBS-related tremor improvement and agreement between objective and subjective measures. **(A)** Total FTM TRS improvement (baseline OFF − mean ON post-operative) stratified by diagnosis category (essential tremor only vs essential tremor with additional diagnosis). No difference in improvement was observed between groups (Welch’s *P* = 0.0866; Wilcoxon rank-sum *P* = 0.0953). **(B)** Total FTM TRS improvement stratified by stimulation laterality relative to the dominant limb (ipsilateral vs contralateral). Improvement did not differ based on whether the dominant side was stimulated (Welch’s *P* = 0.3748; Wilcoxon rank-sum *P* = 0.2928). Box plots show median and interquartile range; whiskers indicate 1.5× interquartile range; individual dots represent single patient measurements. **(C)** Bland–Altman analysis comparing objective clinician-rated tremor severity (FTM TRS motor score) and subjective patient-reported disability (FTM TRS Part C), expressed as a percentage of the maximum possible score for each section. The mean difference (solid line) and limits of agreement (±1.96 SD, dashed lines) indicate substantial inter-individual variability, with subjective ratings generally underestimating objective tremor severity.

**Supplementary Figure 2.**
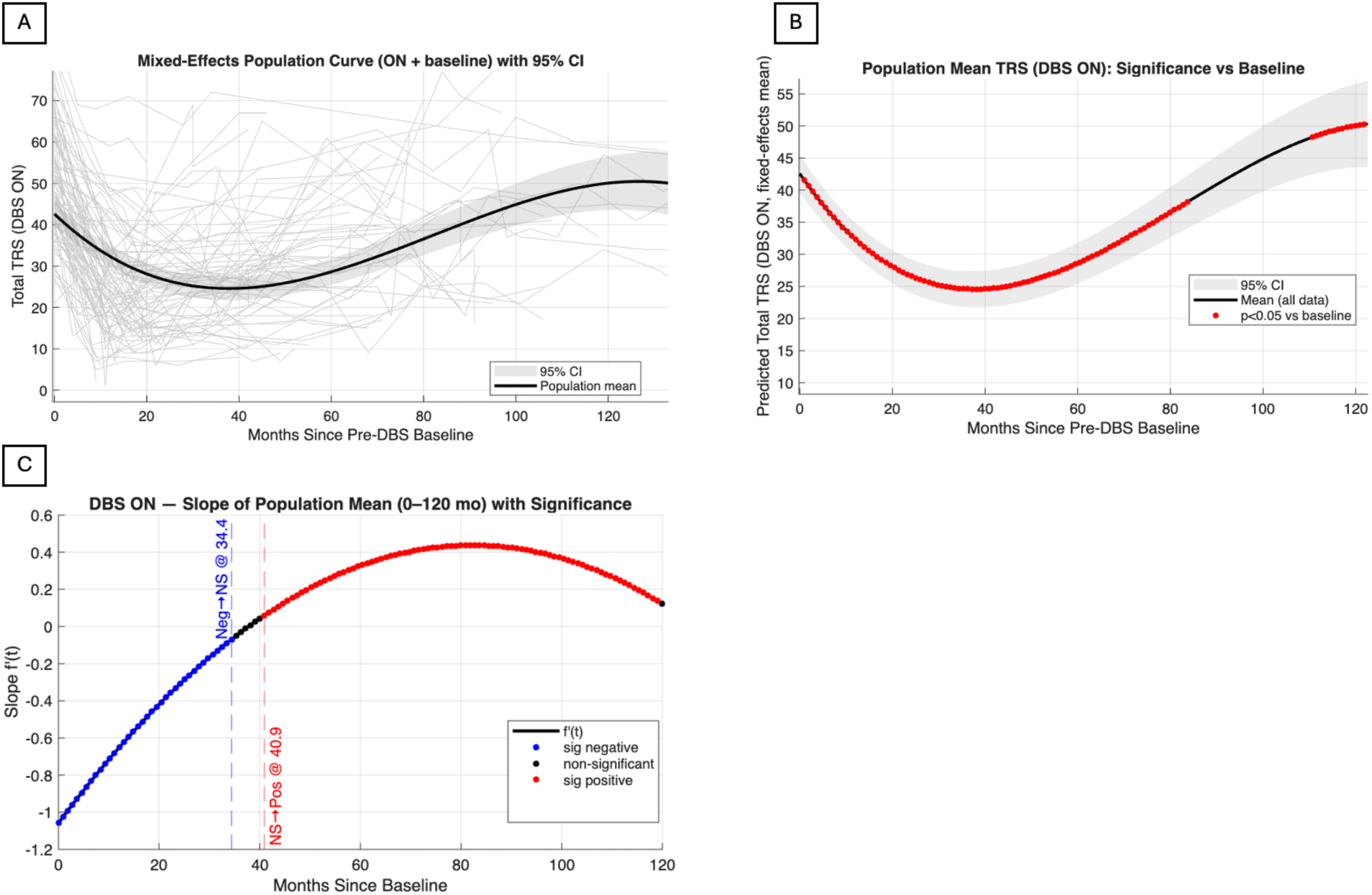
Mixed-effects population trajectories reveal time-dependent improvement and worsening in DBS-ON tremor. **(A)** Individual longitudinal trajectories of total FTM TRS scores in the DBS-ON state, including the pre-operative baseline, shown in light grey. The black curve represents the population-average trajectory estimated using a linear mixed-effects model accounting for repeated measurements within patients. Shaded bands indicate the 95% confidence interval around the population mean. **(B)** Population mean total FTM TRS in the DBS-ON state as a function of time since baseline, estimated using the linear mixed-effects model. Shaded bands indicate the 95% confidence interval. Red points denote time points at which DBS-ON tremor severity differed significantly from baseline (*P* < 0.05). Time points no longer significantly different from baseline indicate a return toward pre-operative tremor severity. **(C)** Time-varying slope of the population mean DBS-ON trajectory over the first 120 months. Blue points indicate intervals with a significantly negative slope (continued improvement), red points indicate a significantly positive slope (worsening), and black points indicate non-significant slope estimates. Vertical dashed lines mark the transitions between significant and non-significant slope regimes.

